# Transitions of care between jail-based medications for opioid use disorder and ongoing treatment in the community: A retrospective cohort study

**DOI:** 10.1101/2023.11.10.23298390

**Authors:** Noa Krawczyk, Sungwoo Lim, Teena Cherian, Keith S. Goldfeld, Monica Katyal, Bianca D. Rivera, Ryan McDonald, Maria Khan, Ellen Wiewel, Sarah Braunstein, Sean M. Murphy, Ali Jalali, Philip J. Jeng, Eric Kutscher, Utsha G. Khatri, Zachary Rosner, William L. Vail, Ross MacDonald, Joshua D. Lee

## Abstract

**Background and Aims:** Offering medications for opioid use disorder (MOUD) in carceral settings can significantly reduce overdose risk. However, it is unknown whether and to what extent individuals in U.S. jail settings continue MOUD once they leave incarceration, and what factors influence treatment continuity.

**Design:** Retrospective cohort study of linked jail-based electronic health records and community OUD treatment claims.

**Setting:** New York City Jail

**Participants:** Incarcerations of individuals with OUD discharged from jail to the community between May 1, 2011 and December 31, 2017

**Comparators:** Receiving vs. not receiving MOUD (methadone or buprenorphine) at the time of release from jail

**Measurements:** We measured continuity of community-based MOUD treatment within one month of release, among those with and without MOUD during release from jail. We tested for effect modification based on MOUD receipt prior to incarceration and assessed factors associated with treatment discontinuation upon re-entry.

**Findings:** Of 28,298 eligible incarcerations, 52.8% received MOUD at release. 30% of incarcerations with MOUD had a community-based MOUD claim within 30 days of release, compared to 7% of incarcerations without MOUD (Risk Ratio: 2.62 (2.44-2.82)). Most (69%) of those with MOUD claims prior to incarceration who received in-jail MOUD continued MOUD in the community, compared to only 9% of those without prior MOUD. Among incarcerations with MOUD at release, those who were younger, Non-Hispanic Black and with no history of MOUD treatment were less likely to continue treatment following release.

**Conclusions:** MOUD maintenance in jail is strongly associated with MOUD continuity in the community upon release. Still, findings highlight a continued gap in MOUD upon-reentry, especially among those who initiate MOUD in jail. In the wake of worsening overdose deaths and troubling disparities, improving continuty of evidence-based care among this population must be an urgent policy priority.

## Introduction

With the worsening U.S. fentanyl epidemic^4^ and deepening disparities in overdose among racial and ethnic minoritized groups,^6,7^ improving initiation and continuity of evidence-based treatments for opioid use disorder (OUD) is an urgent priority.^5^ Methadone and buprenorphine are medications for opioid use disorder (MOUD) that significantly reduce risk of overdose and other negative health outcomes, but are largely under-utilized.^1–3^ Individuals involved in the criminal legal system (CLS) are an important target population that has historically had very low access to MOUD.^8–11^ Risk of overdose is especially elevated in the weeks following release from incarceration,^12,13^ as individuals often lose their opioid tolerance while incarcerated, only to then encounter a dangerous street supply of drugs while facing multiple stressors during re-entry.^14,15^ But offering MOUD treatment during incarceration can mitigate overdose risk upon release:^16–19^ In a recent study of New York City adults leaving NYC jails, for example, those who received MOUD prior to release had nearly half the overdose death rate within a year compared to those who did not.^20^

Despite historical resistance to offering MOUD in the CLS, in recent years, multiple U.S. states have passed laws and regulations mandating that MOUD be made available in carceral settings.^9,21^Critical to expanding jail and prison-based MOUD programs and maintaining their protective effects, however, is ensuring individuals can continue treatment upon release from incarceration. In a cohort study in Australia, receiving MOUD in the 4 weeks post-release from incarceration reduced the hazard of death by 75%.^17^ Randomized trials and pilot studies of jail-based MOUD programs have pointed to the value of jail-based MOUD in increasing the likelihood of engaging in treatment upon release.^22–25^ Still, there remains a dearth of population-based studies that explore whether and to what extent individuals in real-world jail settings continue MOUD once they leave incarceration. More information about the transition between MOUD maintenance during incarceration and community-based treatment, and what factors influence treatment continuity, is key to broadly improving long-term access to MOUD.^9,26–28^

To inform these questions, we analyzed retrospective data of electronic health record (EHR) data from the health provider in the NYC jails – the oldest and one of the most comprehensive correctional MOUD treatment programs in the country^29,30^ – and linked these data to New York State (NYS) Medicaid claims of community-based methadone maintenance treatment (MMT) visits and buprenorphine prescriptions. By doing so, we sought to: 1) assess the relationship between receiving jail-based MOUD and continuing MOUD treatment after release; 2) evaluate whether the association between jail-based MOUD and treatment continuity is moderated by MOUD engagement prior to incarceration; and 3) identify individual sociodemographic, health, substance use, and incarceration-related-legal characteristics associated with treatment discontinuation upon release among those receiving jail-based MOUD.

## Methods

### Setting and Data Sources

This retrospective cohort study focused on incarceration events in the NYC jail with admissions and discharges between May 1, 2011 and December 31, 2017. NYC has one of the largest municipal jail systems in the country: in 2011, the jail had over 85,000 yearly admissions, which were reduced to 56,000 by 2017.^31^ We gathered data using the electronic health records (EHR) of NYC Health + Hospitals Correctional Health Services (CHS), which provides all healthcare, including OUD treatment, social services, and reentry services for all incarcerated individuals at the jails.

Data from CHS were deterministically matched with NYS Medicaid data and probabilistically matched with NYC death certificate data with a matching quality determined to be acceptable (sensitivity = 97%; specificity = 96%). All data were de-identified using a unique patient ID prior to analyses. For full details on the datasets, data selection processes, and probabilistic matching, see Lim et al. (2022).^20^ The study was reviewed and approved by the Institutional Review Boards of the NYC Department of Health and Mental Hygiene (18-106), NYU Langone (i18-00445 and 1811019740), the Weill Cornell Medicine IRB under protocol #1811019740 and the Biomedical Research Alliance of New York (19-PRS-156-419(HHC)), and received certification from the Office for Human Research Protections of the US Department of Health and Human Services.

### Study Population

Since 2011 and prior, all individuals with OUD have been eligible to receive MOUD with methadone or buprenorphine, but individuals who had criminal charges that could result in transfer to prison or other custody (e.g., due to felony charges, parole violations or active warrants, extradition requests) were eligible to receive only acute withdrawal management with MOUD rather than maintenance (largely due to historical lack of MOUD access in NYS prisons). This policy changed in late 2017, making anyone interested in MOUD maintenance eligible for the program. We limited records to incarceration events of adults aged 18-65 with a documented OUD diagnosis based on a comprehensive medical intake evaluation and screening process that takes place upon admission to the jail. As our study focused on community-treatment continuity, we limited our analysis to incarceration events that had a discharge to the community rather than a transfer to prison or another institution, resulting in 70,275 incarceration events.

We then excluded incarceration events with naltrexone (n=1), no MOUD receipt (n=19,645) (i.e. OUD was likely not active), or in which there was a taper at the time of discharge (n=19,504). An MOUD taper potentially indicating ineligibility for maintenance treatment due to legal charge or a short jail stay that precluded initiating a maintenance dose. Finally, we excluded incarceration events after which individuals were either reincarcerated (n=2,776, 9%) or died (n=51, 0.2%) within 30 days post-release, as these events would compete with the ability to enter treatment within the 30-day observation period. Our final analytic dataset included 28,298 incarceration events among 15,609 unique individuals. Of these, we compared 14,948 incarceration events of individuals receiving active MOUD maintenance at the time of release to 13,350 incarceration events of individuals that received MOUD at some point during the jail stay but were not receiving MOUD by the time of release.

### Community Treatment Outcomes

Our main treatment outcome was receipt of MOUD (either methadone or buprenorphine) within 30 days of discharge from jail, as indicated by having at least one Medicaid claim for a community-MMT (methadone maintenance treatment) visit or a community prescription for buprenorphine. Medicaid rate codes and National Drug Codes (NDC) from pharmacy records used to define MMT visits and buprenorphine prescriptions were selected based on standards established in previous protocols utilizing NYS Medicaid data,^32^ and are detailed in Appendix Table 1. In NYS, low-income adults have been eligible for Medicaid since the beginning of the study period, implying that the majority of individuals leaving incarceration in our study cohort would be eligible for Medicaid coverage for OUD treatment and appear in the Medicaid administrative data. Indeed, prior studies of the NYC jail have shown high rates of Medicaid enrollment or history of enrollment among jailed individuals with substance use disorders.^33^

**Table 1.** Characteristics of individuals with incarceration events, by receipt of medications for opioid use disorder (MOUD, i.e. methadone or buprenorphine) within 3 days prior to release, NYC Jail 2011-2017 (n=28,298)

|  | n (%) |  |  |  |
| --- | --- | --- | --- | --- |
|  | Total<br>(N=28,298) | Did not receive<br>MOUD at release<br>(N=13,350) | Received MOUD<br>at release<br>(N=14,948) | Standardized<br>difference |
| **Age (years)** |  |  |  |  |
| 18-24 | 1,424 (5.0) | 978 (7.3) | 446 (3.0) | 0.22 |
| 25-34 | 6,400 (22.6) | 3,207 (24.0) | 3,193 (21.4) |  |
| 35-44 | 7,903 (27.9) | 3,562 (26.7) | 4,341 (29.0) |  |
| 45-54 | 9,512 (33.6) | 4,204 (31.5) | 5,308 (35.5) |  |
| 55-64 | 3,059 (10.8) | 1,399 (10.5) | 1,660 (11.1) |  |
| **Male (vs. female)** | 23,325 (82.5) | 11,885 (89.1) | 11,440 (76.6) | 0.33 |
| **Education Level** |  |  |  | 0.02 |
| Less than High School | 12,178 (44.5) | 5,699 (44.1) | 6,479 (44.9) |  |
| High School Degree or Equivalent | 10,351 (37.8) | 4,864 (37.6) | 5,487 (38.0) |  |
| Some College/Trade School | 4,834 (17.7) | 2,359 (18.3) | 2,475 (17.1) |  |
| **Race/Ethnicity** |  |  |  |  |
| Non-Hispanic, Black | 8,495 (30.1) | 4,111 (30.9) | 4,384 (29.4) | 0.10 |
| Non-Hispanic, White | 6,629 (23.5) | 2,858 (21.5) | 3,771 (25.3) |  |
| Hispanic, Any Race | 12,478 (44.2) | 6,009 (45.1) | 6,469 (43.4) |  |
| Non-Hispanic of another Race | 646 (2.3) | 347 (2.6) | 299 (2.0) |  |
| **Marital Status** |  |  |  |  |
| Single/Never married | 22,162 (78.7) | 10,439 (78.5) | 11,723 (78.8) | 0.04 |
| Married/Partnered | 4,571 (16.2) | 2,206 (16.6) | 2,365 (15.9) |  |
| Divorced/Separated/Widowed | 1,445 (5.1) | 648 (4.9) | 797 (5.4) |  |
| **Unhoused or likely unhoused (vs. stable housing)** | 4,766<br>(16.84%) | 2,009 (15.1) | 2,757 (18.4) | 0.11 |
| **Health-Related** |  |  |  |  |
| History of injection drug use | 12,712 (44.9) | 5,173 (38.8) | 7,539 (50.4) | 0.23 |
| Cocaine Use Disorder | 9,583 (33.9) | 4,032 (30.2) | 5,551 (37.1) | 0.16 |
| Alcohol Use Disorder | 7,007 (24.8) | 3,305 (24.8) | 3,702 (24.8) | 0.00 |
| HIV Diagnosis | 2,240 (7.9) | 947 (7.1) | 1,293 (8.7) | 0.06 |
| HCV Diagnosis | 9,952 (35.2) | 3,719 (27.9) | 6,233 (41.7) | 0.29 |
| Serious Mental Illness | 1,503 (5.3) | 742 (5.6) | 761 (5.1) | 0.02 |
| Any Mental Health Needs | 9,994 (35.3) | 4,829 (36.2) | 5,165 (34.6) | 0.04 |
|  | Total<br>(N=28,298) | Did not receive<br>MOUD at release<br>(N=13,350) | Received MOUD<br>at release<br>(N=14,948) | Standardized<br>difference |
| **Incarceration-Related** |  |  |  |  |
| Prior Incarceration (since 2009) | 23,848 (84.3) | 10,796 (80.9) | 13,052 (87.3) | 0.19 |
| Incarceration lasted >1 month | 13,771 (48.7) | 8,120 (60.8) | 5,651 (37.8) | 0.47 |
| Incarceration related to felony charge | 12,408 (43.9) | 8,272 (62.0) | 4,136 (27.7) | 0.76 |
| Incarceration related to parole violation | 2,087 (7.4) | 1631 (12.2) | 456 (3.1) | 0.35 |
| **Prior Treatment-Related** |  |  |  |  |
| Self-reported opioid use treatment prior to incarceration | 10,528 (37.2) | 2,926 (21.9) | 7,602 (50.9) | 0.63 |
| MOUD claim in two years prior to incarceration | 10,238 (36.2) | 3,393 (25.4) | 6,845 (45.8) | 0.43 |
| MOUD claim in six months prior to incarceration | 7,251 (25.6) | 2,204 (16.5) | 5,047 (33.8) | 0.40 |

### Additional Covariates of Interest

We included additional covariates, recorded in the EHR during incarceration, to examine correlates of community MOUD and to adjust for confounding when examining associations between jail-based MOUD and community MOUD. Sociodemographic data available from the EHR included age at discharge, sex, race/ethnicity, education level, marital status, veteran status, homeless or shelter stay indicated at intake or thereafter, and pregnancy status. We also included substance use and other health indicators, including self-reported injection drug history, cocaine and alcohol use disorder diagnoses, any mental health needs (as indicated based on active jail-based mental health services enrollment), serious mental illness (SMI), and HIV and Hepatitis C (HCV) status. Data on the nature of each incarceration included whether the individual had a prior incarceration (going back to 2009, 2 years prior to the first admission record), whether the incarceration lasted greater than 1 month, and whether the incarceration was for a felony charge or for a prior parole violation at jail admission. We also included CHS covariates on treatment history, including MOUD type received during incarceration (methadone vs. buprenorphine) and self-reported and/or confirmed pre-incarceration OUD treatment. Finally, we used Medicaid claims data from the period prior to incarceration to create two variables on community-MOUD treatment based on whether an individual had at least one claim for MOUD (an MMT visit or buprenorphine prescription) either in the 6 months or 2 years prior to jail admission.

### Statistical Analysis

First, we compared sociodemographic, clinical, incarceration-related, and prior MOUD treatment characteristics between those with “No MOUD at release” and those with “MOUD at release,” using standardized differences (p-values from chi-squared tests would not be informative due to the large sample size). Second, we conducted multivariable robust log linear Poisson regression to estimate risk ratios (RRs) and 95% confidence intervals (CIs) for the relationship between receiving MOUD maintenance while incarcerated and likelihood of having an MOUD claim within 30 days of discharge, accounting for potential confounders of this association. Robust (or modified) Poisson models that use a robust error variance are used widely to estimate risk ratios in cohort studies.^34,35^ We also tested for any effect modification of this relationship based on having had MOUD treatment in the six months prior to incarceration by including an interaction term in our regression model. Third, to understand potential risk factors for discontinuation of MOUD among those who were receiving MOUD maintenance in jail, we limited the sample to those with received MOUD at release and used multivariable robust log linear Poisson regression to assess factors that were associated with likelihood of receiving MOUD within 30 days of discharge. Given the low % of missing data (<3.3% on all covariates), all multivariable regressions were conducted as complete case analyses. Given correlation of multiple incarceration events across unique individuals, we included robust standard errors in all of our regressions. All analyses were conducted in Stata 17.^36^

### Sensitivity Analyses

We conducted multiple sensitivity analyses to ensure our findings were robust to analytic decisions. First, we tested whether findings from our effect modification analysis changed when we tested the effect moderator to be having MOUD claims in the two years rather than six months prior to incarceration. Second, we looked at whether outcomes differed when stratifying by methadone vs. buprenorphine. Third, to ensure there wasn’t significant selection bias given our assessment of community MOUD was limited to Medicaid claims, we assessed whether outcomes differed when we limited the analysis to individuals who had any active enrollment/claim in Medicaid during the month following release, which ensures they were enrolled in Medicaid and any MOUD claims would also appear in the Medicaid dataset. Finally, we assessed whether findings differed when we extended our main outcome to having a claim for MOUD within three months of incarceration release, rather than one month.

## Results

### Characteristics of the study sample

Characteristics of the sample are displayed in Table 1. Among our sample of incarceration events for individuals with OUD, 14,948 (52.8%) received MOUD at release: 13,974 (94%) with MMT and 974 (6%) buprenorphine. Table 1 includes sociodemographic, clinical, incarceration-related, and prior treatment characteristics of the events with MOUD (n=14,948) and those without (n=13,350). Overall, incarceration events were mostly among persons who were male (82%), 35-54 years old (62%), Hispanic (44%) or Non-Hispanic Black (30%). More than one-third self-reported having injected drugs or having mental health needs, 44% of incarcerations were related to a felony, 49% of incarcerations had a duration of a month or longer, and 84% had at least one prior incarceration in NYC in the 2 years prior to the first incarceration event in the study period. Of the sample, 37% self-reported pre-incarceration OUD treatment. Incarceration events with MOUD at release were more likely to be among individuals who were female; who had history of injection, co-occurring cocaine use disorder, or HCV; who were incarcerated <1 month, not facing a felony charge or a parole violation; self-reported prior treatment for OUD, and had a claim for MOUD in the 6 months or 2 years prior to incarceration.

### Association between jail-based MOUD and community MOUD engagement

Among all incarceration events in our sample, 5,373 (19%) received methadone or buprenorphine treatment within one month of discharge as determined by their Medicaid claims (Table 2). Of these, 4,289 (80%) had a claim for a community methadone maintenance treatment (MMT) visit and 1,142 (21%) for a buprenorphine prescription (58 had both an MMT visit and a buprenorphine prescription within the first month).

**Table 2:** Association between receipt of jail-based MOUD at release and community MOUD engagement within one month post-release, stratified by whether individual had an MOUD claim within 6 months prior to incarceration

| Population | No MOUD at release |  | Any MOUD at release |  | Comparison |
| --- | --- | --- | --- | --- | --- |
|  | Total | Received community MOUD within one month post-release | Total | Received community MOUD within one month post-release |  |
|  | N | N (%) | N | N (%) |  |
| **All incarcerations** | 13,350 | 965 (7.2%) | 14,948 | 4,408 (29.5%) | 2.62 (2.44-2.82)<sup>2</sup> |
| **Incarcerations without MOUD claim in six months prior** | 11,154 | 272 (2.4%) | 9,907 | 935 (9.4%) | 5.47 (4.66-6.42)<sup>3</sup> |
| **Incarcerations with MOUD claim in six months prior** | 2,196 | 693 (31.6%) | 5,041 | 3,473 (68.9%) | 2.01 (1.87-2.15)<sup>3</sup> |

Notes:
Adjusted risk ratios (aRR) were calculated using robust log linear Poisson regression. All models account for clustering by person and complete-case analysis and exclude individuals who died within 30 days (n=51) or were re-incarcerated within 30 days (n=2,776)
<sup>1</sup>All multivariable models account for age, sex, education level, race/ethnicity, marital status, history of injection drug use, serious mental illness, cocaine use disorder, alcohol use disorder, HIV diagnosis, HCV diagnosis, unhoused/unstably housed, mental health needs, prior incarceration (since 2009), incarceration lasted >1 month, incarceration related to felony charge, incarceration related to parole violation, self-reported opioid use disorder treatment prior to incarceration, unhoused/unstably housed, mental health needs.
<sup>2</sup>Multivariable all incarcerations model additionally adjusts for prior MOUD treatment within 6 months of incarceration.
<sup>3</sup>Interaction coefficient between past-6-months MOUD and MOUD was significant at p<0.05 level (0.56 p=<0.001)

As shown in Table 2, among incarcerations with MOUD at release, 30% had a subsequent community-based MOUD claim within 30 days, while only 7% of incarcerations without MOUD at release had a community-based MOUD claim within 30 days. After adjusting for potential confounders as well as prior treatment history, individuals with MOUD were 2.62 times as likely to enter MOUD within one month (95% Confidence Interval [CI]: 2.44-2.82).

A majority of those who had in-jail MOUD and a prior MOUD claim continued MOUD in the community (69%), while only 9% of individual events with new initiation of MOUD in-jail continued MOUD in the community (Table 2). In both groups, having MOUD at release was associated with a significantly increased probability of entering MOUD within one month of discharge, with an even larger association for those without prior MOUD (Adjusted Risk Ratio (aRR): 5.47 (95% CI: 4.66-6.42)) than for those with prior MOUD (aRR: 2.01 (95% CI: 1.87-2.15)). Prior MOUD use significantly modified the effect of MOUD at release (interaction term p<0.001)

### Correlates of community MOUD continuity among those receiving MOUD at release

When we limited the sample to incarceration events in which individuals did receive MOUD at release, multiple characteristics were independently associated with subsequent community MOUD (Figure 1): having an MOUD treatment claim in the six months prior to the index incarceration (aRR: 6.76 (95% CI: 6.30-7.27), older age (e.g., 55-64 vs. 18-24, aRR: 1.37 (95% CI: 1.13-1.64)), and being male (aRR: 1.10 (95% CI: 1.04-1.17)). Non-Hispanic Black individuals were less likely to continue MOUD compared to Non-Hispanic white individuals (aRR: 0.89 (95% CI: 0.83-0.95)), while Hispanic individuals and those of another race/ethnicity had a similar likelihood to continue MOUD compared to Non-Hispanic white individuals.

**Figure 1:**
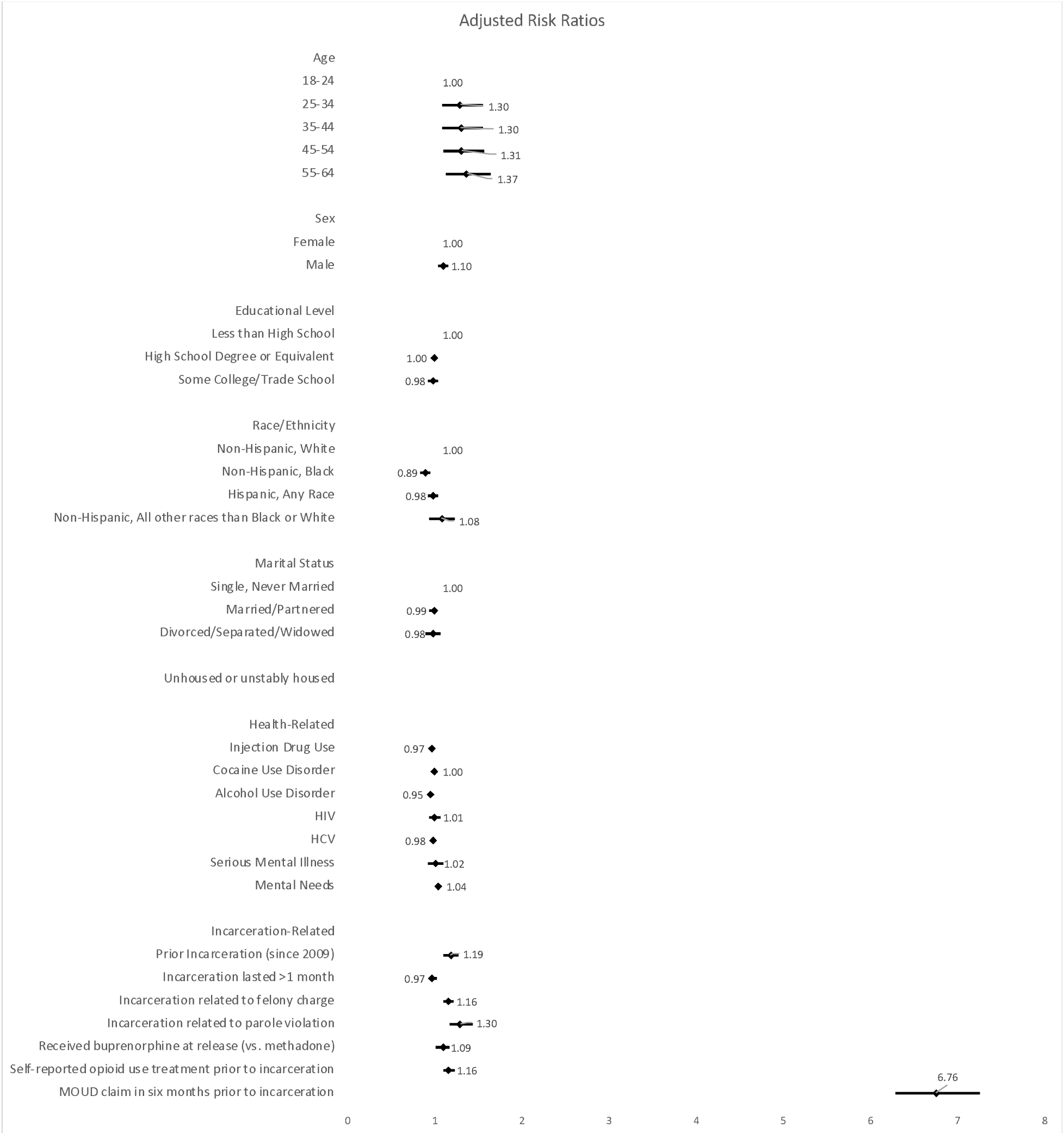
Factors associated with MOUD engagement at one month among people who receives MOUD at release based on multivariable analysis (N=14,948) Note: Adjusted and unadjusted estimates and confidence intervals are available in Appendix Table 6

Multiple characteristics of the incarceration itself were associated with MOUD continuity, including having had a prior incarceration (aRR: 1.19 (95% CI: 1.10-1.27)), felony charge (aRR: 1.16 (95% CI: 1.10-1.22)) or parole violation (aRR: 1.30 (95% CI: 1.17-1.43)). Those who received buprenorphine in jail had a slightly greater likelihood of continuing MOUD compared to those who received methadone (aRR: 1.09 (95% CI: 1.02-1.18)) as did those who reported being in treatment prior to incarceration compared to those who did not (aRR: 1.16 (95% CI: 1.10-1.24)). None of the health-related factors we assessed were associated with treatment continuation, nor was housing stability or marital status.

### Sensitivity Analyses

Results of sensitivity analyses can be found in Appendix Tables 2-6. First, when we tested whether the main association between in-jail MOUD and community MOUD was moderated by prior MOUD treatment and expanded prior MOUD treatment to include those with any prior two-year MOUD claims (vs. six-months only), findings did not qualitatively change, and the interaction between prior MOUD claims and MOUD at release was still highly significant (Appendix Table 2). Second, a strong association between in-jail MOUD and MOUD continuity within the 1^st^ month of discharge continued when we analyzed data separately for those who received methadone vs. buprenorphine, with in-jail MMT having a particularly strong association with receiving community-based MMT relative to those not receiving MMT in jail (aRR: 3.98 (95% CI: 3.60-4.40)) (Appendix Table 3). We also found that results did not qualitatively change when we limited the dataset to individuals with active Medicaid enrollment upon release (Appendix Table 4), indicating findings were not likely a result of disproportionate Medicaid eligibility among the MOUD vs. non-MOUD groups. Finally, the impact of in-jail MOUD on community-based MOUD slightly decreased but was still strong (aRR: 2.11 (2.00-2.23)) when we extended the main outcome to having a claim for MOUD within three months of incarceration discharge, rather than one month (Appendix Table 5).

## Discussion

The current study provides a unique opportunity to observe real-world patterns and correlates of MOUD continuity after release from incarceration that is rarely available in US population-based samples. Study findings corroborate prior clinical trial and pilot study results^22–24,37^ indicating that jail-based MOUD is strongly associated with MOUD continuity upon release. This effect was even larger for those initiating MOUD without prior treatment, pointing to the importance of offering jail-based MOUD initiation in addition to maintenance. Continuity of MOUD in the post-incarceration period is critical to sustaining improved health outcomes offered by these treatments, including reducing overdose risk and healthcare costs.^3,38^ Another recent analysis from our team using these data found that community utilization of MOUD significantly mediated the effect of in-jail MOUD on mortality outcomes within a year of jail release.^39^

Despite improved MOUD linkage outcomes among people receiving MOUD in jail, overall rates of transition to community treatment among this group were still low, below 30%. The proportion of individuals entering community MOUD was not substantially larger by three months post-release (35%), indicating MOUD engagement rates are unlikely to rise over time. Community re-entry is known to be a high-risk time, and these low rates of community linkage to care are in line with those reported for other serious conditions such as HIV.^40^ Still, this low rate is concerning given NYC jail’s MOUD program is one of the most comprehensive in the country, offering both initiation and maintenance for both methadone and buprenorphine. Many U.S. jails do not routinely offer universal access to MOUD or do so on a limited basis, such as for those already in MOUD treatment or special populations like pregnant women, or prioritize naltrexone over opiate agonist medications.^9,41,42^ In addition, few other cities and counties have access to as many community MOUD providers as are available in NYC,^43,44^ with lack of community MOUD availability often cited by jail leaders as a challenge for offering and linking incarcerated individuals to MOUD.^26^

The low rate of MOUD continuity observed in our data was overwhelmingly driven by low (9%) enrollment after release by people initiated on MOUD in jail, whilst a majority (69%) of those who had treatment prior to incarceration and received MOUD in jail did go on to continue treatment in the community. This was despite the fact that reeiving MOUD in jail at release, regardless of prior treatment, was still a strong predictor of MOUD following discharge. Therefore, better incentives and opportunities for engagement in care post-release are needed for individuals initiated onto MOUD during incarceration. The same is true for younger adults, who regardless of pre-incarceration history of MOUD, had significantly worse MOUD continuity post-release than older persons, and for whom MOUD engagement is known to be particularly challenging.^45,46^ ^47,48^

Non-Hispanic Black individuals in our sample were also less likely to continue MOUD upon release. Prior studies have found mixed findings on racial differences in MOUD continuity, with a meta-analysis of retention in MOUD finding no differences across race/ethnicity groups, yet a recent study finding lower MOUD retention rates among Non-Hispanic Black individuals receiving buprenorphine.^49^ Our study highlights a need for greater efforts to understand and address structural barriers to care and treatment continuity, especially among minoritized groups, for whom racist drug war and policing practices,^50^ as well as a long history of discrimination in substance treatment and other healthcare settings, have resulted in disproportionate criminalization, marginalization, and, increasingly, worse overdose outcomes.^7,50–52^

Challenges for continuing MOUD after incarceration, particularly among those initiating care in jail, may reflect the chaotic nature of jail-reentry (e.g. unplanned discharges, little opportunity to discuss discharge plans) and its synergistic effects with larger barriers to MOUD utilization in the community. A recent national analysis estimated that only 13% of those with OUD receive MOUD.^53^ Those with criminal-legal involvement additionally experience a range of social, medical, and structural barriers to these treatments.^54–56^ Expanding the availability of providers that offer MOUD in low-threshold settings that can cater to the needs of this population during the chaotic period of re-entry might be particularly important. Examples of such programs include mobile van settings^57^ or telehealth buprenorphine clinics, which have been shown to be successful in catering to incarcerated individuals at reentry,^58,59^ as well as the Transitions Clinic Network, a consortium of primary care clinics that aims to increase access to health care services among people recently released from incarceration.^60^ Recent trials have also shown the potential promise of injectable forms of buprenorphine in facilitating treatment continuity following discharge,^61–63^ but uptake of these treatments in carceral settings has been slow.

Interestingly, those in our sample who received buprenorphine (vs. methadone) in jail were more likely to continue MOUD, which is consistent with findings from a prior clinical trial in NYC.^64^ Improved continuity of buprenorphine may relate in part to greater facility in accessing buprenorphine via office-based prescribing, compared to methadone, which can only be dispensed at highly-regulated and often burdensome opioid treatment programs (OTPs). Continued efforts to increase COVID-19 flexibilities for methadone take-homes and facilitating the initiation of buprenorphine via telehealth, which has been found to facilitate initiation and retention in MOUD, are strategies that can help facilitate long-term outcomes in these treatments.^65,66^ Expanding methadone to other treatment settings, including mobile health vans (which have recently launched in NY)^67,68^ and office-based practices as is done in many non-US settings, ^69^and which has been proposed by recent legislation, may also improve MMT access and continuity. Importantly, ensuring connections to harm reduction programs is also critical for ensuring reduced overdose among this population. Expanding alternatives to incarceration and reducing incarceration itself remains a vital goal as it is known to highly destabilize multiple factors that promote stability such as employment, housing, and family connections.^70^ Finally, in addition to expanding the number of settings where evidence-based services can be obtained, ensuring training of providers in person-centered, non-stigmatizing care for people with a history of incarceration would be critical to improving health outcomes among this group.

Lastly, our study found that even in the NYC jail system, which has a robust MOUD program, only half of the OUD patient sample was receiving MOUD at the time of release. This is explained mostly by historical limit eligibility for MOUD based on charges (reflecting the historical unavailability of MOUD in NYS prisons), a policy that was later changed in 2017. Indeed, those with a felony charge and parole violation or with longer incarceration periods in our sample were least likely to have MOUD at release. This points to the importance of making MOUD available in all prison systems to ensure care continuity and access to evidence-based treatment regardless of charge or disposition. Unfortunately, most prison systems across the U.S. still do not offer MOUD.^71^ Those not receiving MOUD maintenance were also more likely to be younger, male, not be an injection drug user and not have co-occurring disorders such as cocaine use, HIV or HCV, potentially indicating lower severity of OUD for which incarcerated individuals or health care providers in jails may have decided against MOUD. Greater efforts to understand decision making around in-jail MOUD, including the role of stigma, mistrust, and preferences among patients even when MOUD is available,^72^ is important for informing continuous efforts to offer evidence-based treatment in these settings and upon release, particularly as efforts grow across the U.S. to increase access to MOUD in both jail and prison settings.^9^

This study is subject to multiple limitations. First, our outcomes data on community-MOUD was only limited to observable Medicaid claims, and individuals who received MOUD covered by other insurance types or private pay may have been missed by this analysis. However, given that the great majority of individuals leaving NYC jail are Medicaid-eligible, and that our sensitivity analyses restricting to those actively enrolled in Medicaid did not portray different findings, this limitation does not likely introduce significant bias. Second, our outcome variable focused on whether individuals received any claim for MOUD, and we did not measure retention or differentiate between individuals who continued or discontinued MOUD after the first claim. Third, our analytic sample of individuals leaving incarceration excluded individuals who were receiving MOUD for withdrawal management only (as opposed to MOUD maintenance treatment). Thus, we are not able to draw conclusions about the impact of MOUD used for withdrawal management (e.g. detox) on subsequent treatment outcomes. Lastly, given our sample was uniquely situated in the NYC jails, which have a robust MOUD infrastructure and a wide network of MOUD providers, these findings may not generalize to other U.S. settings which generally have low access to MOUD in correctional facilities and where community-based MOUD providers may not be as readily present.

## Conclusions

MOUD maintenance while in jail is strongly associated with going on to receive MOUD in the community upon release, regardless of prior history of MOUD. MOUD access in jail mitigated discontinuity for those with previous treatment experience and resulted in better treatment continuity upon return to the community among those initiated on MOUD during incarceration. Still, findings highlight a continued gap in MOUD continuity upon re-entry even among those who do receive MOUD in-jail, especially among certain groups already experiencing barriers to community treatment: persons who are younger, Non-Hispanic Black, or newly initiated on MOUD while incarcerated. Findings support the dire need to invest in re-entry programs and low-barrier treatment models to enable MOUD continuity among the highest-risk groups, while emphasizing the need for alternative pathways to engage individuals in treatment and harm reduction services without the criminal justice system as an intermediary. In the wake of worsening overdose deaths which reveal troubling disparities, improving access to evidence-based care among this population must be an urgent public health and policy priority.

## Data Availability

All data produced in the present study are protected and not publicly available or sharable due to sensitive nature of data (EHR, Medicaid claims, mortality records)

## Appendix Tables

**Appendix Table 1:**
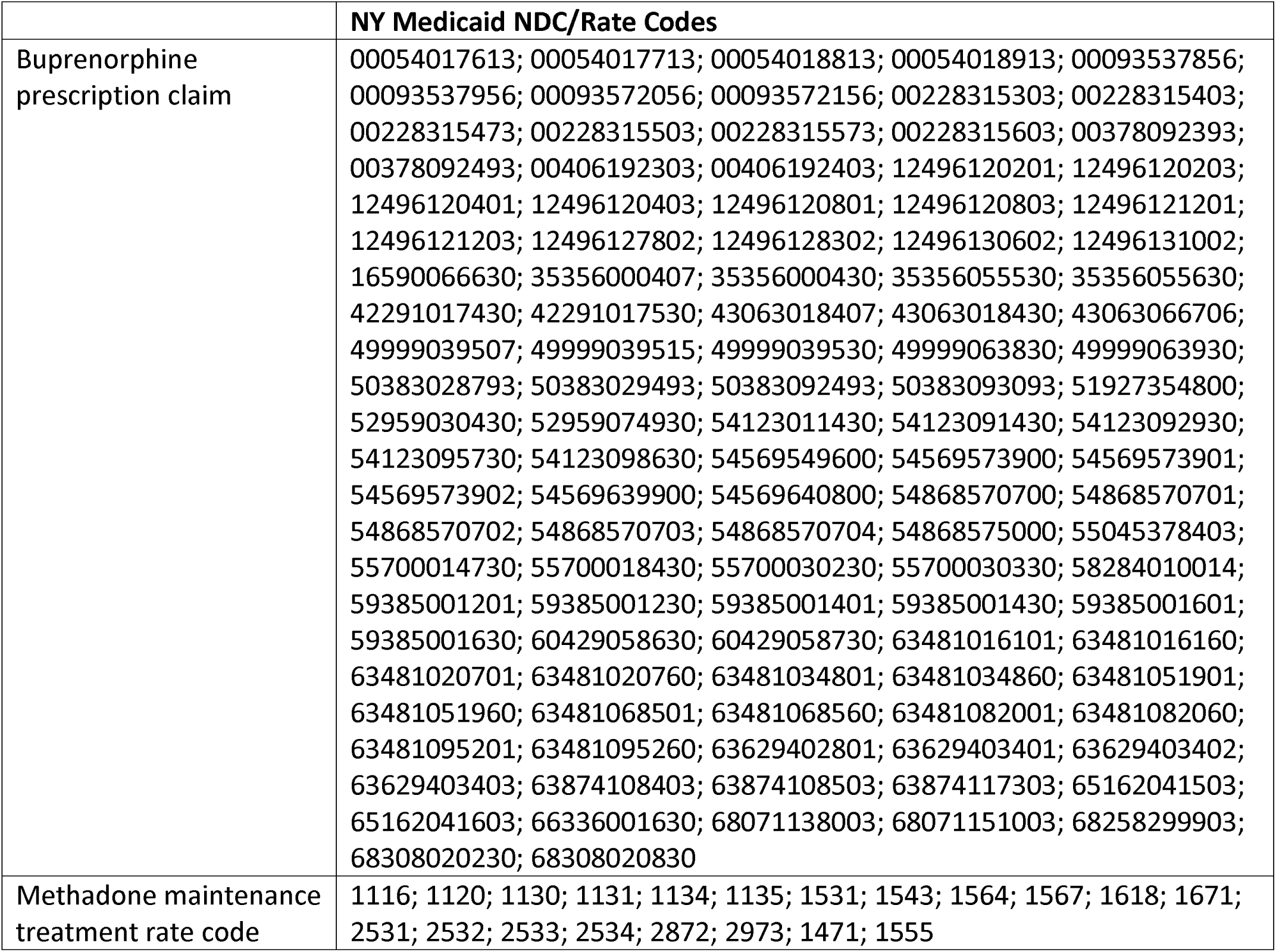
Rate codes and definitions used to define opioid treatment program visits and buprenorphine prescriptions in Medicaid data.

**Appendix Table 2:** Association between receipt of jail-based MOUD and community MOUD engagement within one month post-release, stratified by whether individual had MOUD claim within two years prior to incarceration.

| Population | No MOUD at release |  | Any MOUD at release |  | Comparison |
| --- | --- | --- | --- | --- | --- |
|  | Total | Received community MOUD within one month post-release | Total | Received community MOUD within one month post-release |  |
|  | N | N (%) | N | N (%) |  |
| **Incarcerations without MOUD claim in two years prior** | 8,113 | 166 (1.67%) | 9,965 | 585 (7.21%) | **6.97 (5.68-8.55)<sup>2</sup>** |
| **Incarcerations with MOUD claim in two years prior** | 3,385 | 799 (23.60%) | 6,835 | 3,823 (55.93%) | **2.20 (2.05-2.36)<sup>2</sup>** |

Notes:
Adjusted risk ratios (aRR) were calculated using robust log linear Poisson regression. All models account for clustering by person and complete-case analysis and exclude individuals who died within 30 days (n=51) or were re-incarcerated within 30 days (n=2,776)
<sup>1</sup>All multivariable models account for age, gender, education level, race/ethnicity, marital status, history of injection drug use, serious mental illness, cocaine use disorder, alcohol use disorder, HIV diagnosis, HCV diagnosis, unhoused/unstably housed, mental health needs, prior incarceration (since 2009), incarceration lasted >1 month, incarceration related to felony charge, incarceration related to parole violation, self-reported opioid use disorder treatment prior to incarceration, unhoused/unstably housed, mental health needs.
<sup>2</sup>Interaction coefficient between past-6-months MOUD and MOUD at release was significant at the p<0.05 level (0.52 p=<0.001)

**Appendix Table 3:** Association between receipt of jail-based MOUD and community MOUD engagement one month post-release; analyzed separately for methadone and buprenorphine.

3A. Methadone maintenance treatment (MMT)
| Population | No MOUD<sup>1</sup> at release |  | Any MMT at release |  | Comparison |
| --- | --- | --- | --- | --- | --- |
|  | Total | Received community MMT within one month post-release | Total | Received community MMT within one month post-release |  |
|  | N | N (%) | N | N (%) |  |
| All incarcerations<sup>2</sup> | 13,350 | 499 (3.74%) | 13,974 | 3,733 (26.71%) | **3.98 (3.60-4.40)** |
| Incarcerations without MOUD claim in six months prior | 11,154 | 170 (1.42%) | 9,947 | 913 (9.18%) | **9.65 (7.96-11.69)** |
| Incarcerations with MOUD claim in six months prior | 1,364 | 329 (24.12%) | 4,027 | 2,820 (70.03%) | **2.56 (2.32-2.83)** |

3B. Buprenorphine treatment
| Population | No MOUD<sup>1</sup> at release |  | Any Buprenorphine at release |  | Comparison |
| --- | --- | --- | --- | --- | --- |
|  | Total | Received community bup. within one month post-release | Total | Received community bup. within one month post-release |  |
|  | N | N (%) | N | N (%) |  |
| All incarcerations<sup>2</sup> | 13,350 | 478 (3.58%) | 974 | 398 (40.86%) | **2.36 (2.05-2.72)** |
| Incarcerations without MOUD claim in six months prior | 12,404 | 173 (1.39%) | 484 | 49 (10.12%) | **6.56 (4.64-9.27)** |
| Incarcerations with MOUD claim in six months prior | 946 | 305 (32.24%) | 490 | 349 (71.22%) | **1.84 (1.64-2.07)** |

Notes:
Adjusted risk ratios (aRR) were calculated using robust log linear Poisson regression. All models account for clustering by person and complete-case analysis and exclude individuals who died within 30 days (n=51) or were re-incarcerated within 30 days (n=2,776)
<sup>1</sup>All multivariable models account for age, gender, education level, race/ethnicity, marital status, history of injection drug use, serious mental illness, cocaine use disorder, alcohol use disorder, HIV diagnosis, HCV diagnosis, unhoused/unstably housed, mental health needs, prior incarceration (since 2009), incarceration lasted >1 month, incarceration related to felony charge, incarceration related to parole violation, self-reported opioid use disorder treatment prior to incarceration, unhoused/unstably housed, mental health needs.
<sup>2</sup>Multivariable all incarcerations model additionally adjusts for prior MOUD treatment within 6 months of incarceration.

**Appendix Table 4:** Association between receipt of jail-based MOUD and community MOUD engagement one month post-release, limited to individuals with active enrollment in Medicaid within one month of release.

| Population | No MOUD<sup>1</sup> at release |  | Any MOUD at release |  | Comparison |
| --- | --- | --- | --- | --- | --- |
|  | Total | Received community MOUD within one month post-release | Total | Received community MOUD within one month post-release |  |
|  | N | N (%) | N | N (%) |  |
| **All incarcerations<sup>2</sup>** | 8,744 | 961 (10.99%) | 10,848 | 4,402 (34.45%) | **2.59 (2.42-2.78)** |
| **Incarcerations without MOUD claim in six months prior** | 6,571 | 272 (4.14%) | 5,834 | 934 (16.01%) | **5.13 (4.39-5.99)** |
| **Incarcerations with MOUD claim in six months prior** | 2,173 | 689 (31.71%) | 5,014 | 3,468 (69.17%) | **2.01 (1.87-2.15)** |

**Notes:**
Adjusted risk ratios (aRR) were calculated using robust log linear Poisson regression. All models account for clustering by person and complete-case analysis and exclude individuals who died within 30 days (n=51) or were re-incarcerated within 30 days (n=2,776)
<sup>1</sup>All multivariable models account for age, gender, education level, race/ethnicity, marital status, history of injection drug use, serious mental illness, cocaine use disorder, alcohol use disorder, HIV diagnosis, HCV diagnosis, unhoused/unstably housed, mental health needs, prior incarceration (since 2009), incarceration lasted >1 month, incarceration related to felony charge, incarceration related to parole violation, self-reported opioid use disorder treatment prior to incarceration, unhoused/unstably housed, mental health needs.
<sup>2</sup>Multivariable all incarcerations model additionally adjusts for prior MOUD treatment within 6 months of incarceration.

**Appendix Table 5:** Association between receipt of jail-based MOUD and community MOUD engagement within 3 months post-release, stratified by whether individual had an MOUD claim within 6 months prior to incarceration.

| Population | No MOUD<sup>1</sup> at release |  | Any MOUD at release |  | Comparison |
| --- | --- | --- | --- | --- | --- |
|  | Total | Received community MOUD within three months post-release | Total | Received community MOUD within three months post-release |  |
|  | N | N (%) | N | N (%) |  |
| All incarcerations<sup>2</sup> | 13,350 | 1,513 (11.33%) | 14,948 | 5,150 (34.45%) | **2.11 (2.00-2.23)** |
| Incarcerations without MOUD claim in six months prior | 11,154 | 561 (5.03%) | 9,907 | 1,316 (13.28%) | **3.37 (3.00-3.78)** |
| Incarcerations with MOUD claim in six months prior | 2,196 | 952 (43.35%) | 5,041 | 3,834 (76.06%) | **1.68 (1.59-1.78)** |

Notes:
Adjusted risk ratios (aRR) were calculated using robust log linear Poisson regression. All models account for clustering by person and complete-case analysis and exclude individuals who died within 30 days (n=51) or were re-incarcerated within 30 days (n=2,776)
<sup>1</sup>All multivariable models account for age, gender, education level, race/ethnicity, marital status, history of injection drug use, serious mental illness, cocaine use disorder, alcohol use disorder, HIV diagnosis, HCV diagnosis, unhoused/unstably housed, mental health needs, prior incarceration (since 2009), incarceration lasted >1 month, incarceration related to felony charge, incarceration related to parole violation, self-reported opioid use disorder treatment prior to incarceration, unhoused/unstably housed, mental health needs.
<sup>2</sup>Multivariable all incarcerations model additionally adjusts for prior MOUD treatment within 6 months of incarceration.

**Appendix Table 6:** Factors associated with MOUD engagement at one month among people who receives MOUD at release (N=14,948)

|  | Unadjusted Risk Ratio (95% CI) | Adjusted Risk Ratio (95% CI) |
| --- | --- | --- |
| **Age** |  |  |
| 18-24 | 1.00 | 1.00 |
| 25-34 | **1.49 (1.18, 1.89)** | **1.30 (1.08, 1.55)** |
| 35-44 | **1.66 (1.31, 2.10)** | **1.30 (1.09, 1.55)** |
| 45-54 | **1.62 (1.28, 2.05)** | **1.31 (1.09, 1.57)** |
| 55-64 | **1.79 (1.39, 2.29)** | **1.37 (1.13, 1.64)** |
| **Gender** |  |  |
| Female | 1.00 | 1.00 |
| Male | **1.26 (1.15, 1.37)** | **1.10 (1.04, 1.17)** |
| **Educational Level** |  |  |
| Less than High School | 1.00 | 1.00 |
| High School Degree or Equivalent | 0.98 (0.92, 1.05) | 1.00 (0.95, 1.04) |
| Some College/Trade School | 0.91 (0.83, 1.00) | 0.98 (0.92, 1.04) |
| **Race/Ethnicity** |  |  |
| Non-Hispanic, White | 1.00 | 1.00 |
| Non-Hispanic, Black | **0.82 (0.75, 0.90)** | **0.89 (0.83, 0.95)** |
| Hispanic, Any Race | **1.09 (1.01, 1.18)** | 0.98 (0.93, 1.04) |
| Non-Hispanic, All other races than Black or White | 1.05 (0.87, 1.28) | 1.08 (0.94, 1.23) |
| **Marital Status** |  |  |
| Single, Never Married | 1.00 | 1.00 |
| Married/Partnered | 1.04 (0.97, 1.13) | 0.99 (0.94, 1.05) |
| Divorced/Separated/Widowed | 1.08 (0.95, 1.22) | 0.98 (0.90, 1.07) |
| Unhoused or unstably housed | **1.11 (1.04, 1.19)** | 1.04 (0.99, 1.09) |
| **Health-Related** |  |  |
| Injection Drug Use | 1.05 (0.99, 1.12) | 0.97 (0.92, 1.01) |
| Cocaine Use Disorder | **0.90 (0.85, 0.95)** | 1.00 (0.96, 1.05) |
| Alcohol Use Disorder | **0.82 (0.76, 0.88)** | 0.95 (0.91, 1.00) |
| HIV | 1.10 (0.99, 1.22) | 1.01 (0.94, 1.08) |
| HCV | **1.13 (1.06, 1.21)** | 0.98 (0.94, 1.03) |
| Serious Mental Illness | **1.12 (1.00, 1.26)** | 1.02 (0.93, 1.11) |
| Mental Needs | **1.16 (1.10, 1.23)** | 1.04 (0.99, 1.09) |
| **Incarceration-Related** |  |  |
| Prior Incarceration (since 2009) | **1.46 (1.33, 1.61)** | **1.19 (1.10, 1.27)** |
| Incarceration lasted >1 month | **1.15 (1.09, 1.21)** | 0.97 (0.93, 1.02) |
| Incarceration related to felony charge | **1.31 (1.24, 1.39)** | **1.16 (1.10, 1.22)** |
| Incarceration related to parole violation | **1.81 (1.65, 2.00)** | **1.30 (1.17, 1.43)** |
| Received buprenorphine at release (vs. methadone) | **1.62 (1.49, 1.76)** | **1.09 (1.02, 1.18)** |
| Self-reported opioid use treatment prior to incarceration | **1.99 (1.87, 2.11)** | **1.16 (1.10, 1.24)** |
| MOUD claim in six months prior to incarceration | **7.32 (6.83, 7.83)** | **6.76 (6.29, 7.26)** |

